# Early Experience With Neutralizing Monoclonal Antibody Therapy For COVID-19

**DOI:** 10.1101/2021.04.09.21255219

**Authors:** Mark Jarrett, Warren B. Licht, Kevin Bock, Zenobia Brown, Jamie S. Hirsch, Kevin Coppa, Rajdeep Brar, Steven Bello, Ira S. Nash

## Abstract

**Importance:** Neutralizing monoclonal antibody (MAB) therapies may benefit patients with mild to moderate COVID-19 at high risk for progressing to severe COVID-19 and/or hospitalization. Studies documenting approaches to deliver MAB infusions as well as demonstrating their efficacy are lacking.

**Objective:** We describe our experience and patient outcomes of almost 3,000 patients who received MAB infusion therapy at Northwell Health, a large integrated health care system in New York.

**Design, Setting, and Participants:** This is a descriptive study of adult patients who received MAB therapy between November 20, 2020, to January 31, 2021, and a retrospective cohort survival analysis comparing patients who received MAB therapy prior to admission versus those who did not. A multivariable Cox model with inverse probability weighting according to the propensity score including covariates (sociodemographic, comorbidities, and presenting vital signs) was used.

**Main outcomes and measures:** The primary outcome was in-hospital mortality; additional evaluations included ED utilization and hospitalization within 28 days of a positive COVID-19 test for patients who received MAB therapy.

**Results:** During the study period, 2818 adult patients received MAB infusion. Following therapy and within 28 days of COVID-19 test, 123 patients (4.4%) presented to the ED and were released and 145 patients (5.1%) were hospitalized. These 145 patients were compared with 200 controls who were eligible for but did not receive MAB therapy, and were hospitalized. In the MAB group, 16 (11%) patients met the primary outcome of in-hospital mortality, versus 21 (10.5%) in the control group. In an unadjusted Cox model, the hazard ratio (HR) for time to in-hospital mortality for the MAB group was 1.38 (95% confidence interval [95% CI] 0.696-2.719). Models adjusting for demographics (HR 1.1, 95% CI 0.53-2.23), demographics and Charlson Comorbidity Index (CCI) (HR 1.22, 95% CI 0.573-2.59), and with inverse probability weighting according to propensity scores (HR 1.19, 95% CI 0.619-2.29) did not demonstrate significance. The hospitalization rate was 4.4% for patients who received MAB therapy within 0-4 days, 5% within 5-7 days, and 6.1% within ≥8 days of symptom onset (p-value = 0.15).

**Conclusions and relevance:** Establishing the capability to provide neutralizing MAB infusion therapy requires significant planning and coordination. While this therapy may be an important treatment option for early mild to moderate COVID-19 in high-risk patients, further investigations are needed to define the optimal timing of MAB treatment in order to reduce hospitalization and mortality.

## INTRODUCTION

In November 2020, the Federal Drug Administration (FDA) issued an Emergency Use Authorization (EUA) for the neutralizing monoclonal antibody (MAB) infusions bamlanivimab and casirivimab/imdevimab for treatment of early mild to moderate SARS-CoV-2 infection in patients at high risk for progressing to severe Coronavirus Disease 2019 (COVID-19) and/or hospitalization.^1^ Bamlanivimab has been found to decrease viral load at 11 days, and exploratory analysis of COVID-19 related hospitalization suggested a decrease in hospitalization rate from 6.3% to 1.6%.^2^ Additional studies of bamlanivimab in combination with etesevimab also found reductions in viral load, and similarly found a reduction in hospitalization, although the latter was not the primary outcome.^3^ Most recently, bamlanivimab co-administered with remdesivir did not demonstrate efficacy among hospitalized patients with COVID-19 without end-organ failure.^4^ To date, published data on the effectiveness of these therapies is mixed, and the NIH correspondingly notes that data are insufficient to recommend for or against the use of MAB therapy for ambulatory patients.^5^

Given the operational complexity and uncertain clinical effectiveness of setting up a MAB infusion program, widespread use has been slow across the United States.^6^ Potential barriers to implementation include staffing challenges during disease resurgence, the necessity to provide infusions in a COVID-19 contained environment, transportation of underserved and elderly patients to infusion centers, and the need to obtain timely referrals from providers.^7^ Mobile units have shown to be successful in a small study,^8^ although the ability to scale this solution appears limited. The Mayo Clinic recently reported their implementation of a program across multiple facilities in different states, culminating in over 4,000 doses delivered.^9^

Northwell Health, a 23-hospital integrated health care system in metropolitan New York, established outpatient infusion centers based on their experience with the Spring 2020 surge,^10^ which stretched inpatient capacity. At the peak of the early surge, Northwell had more than 3400 COVID-19 inpatients, with over 800 receiving invasive mechanical ventilation. With the goal of reducing hospitalizations, ICU admissions, and deaths during the Fall and Winter 2020 rise in cases, Northwell rapidly scaled a MAB infusion program. We reviewed our initial experience in using MAB therapy, and describe the outcomes of almost 3,000 patients who received this outpatient infusion therapy, the largest cohort with outcomes published to date.

## METHODS

This was a retrospective study of a large integrated health care system, with 23 hospitals and over 800 ambulatory locations. Data for this study were obtained from the enterprise inpatient and outpatient electronic health record (EHR; Sunrise Clinical Manager and Touchworks, respectively [Allscripts, Chicago, IL]), our Health Information Exchange (Healthshare [Intersystems, New York, NY]), and our locally developed population health management tool (CareTool Listapp [Northwell Health, Lake Success, NY]).

### Monoclonal Antibody Infusion Eligibility Criteria

Eligibility to receive MAB therapy as directed by the FDA EUA is limited to patients with a positive direct viral test for SARS-CoV-2 within 10 days of symptom onset. Patients must be ≥12 years of age, weigh at least 40 kg, and at high risk for progressing to severe COVID-19 or hospitalization. High risk is defined as having one of the following conditions: age ≥65 years, obesity (body mass index [BMI] ≥35 kg/m^2^), diabetes mellitus (DM), chronic kidney disease (CKD), immunosuppressive disease or currently receiving immunosuppressive treatment. Patients 55-64 years old who have cardiovascular disease, hypertension, chronic obstructive pulmonary disease (COPD), or chronic respiratory disease also are eligible. Pediatric patients ages 12-17 with one of the following conditions are also eligible: BMI ≥85th percentile for age and gender, sickle cell disease, congenital or acquired heart disease, neurodevelopmental disorders, a medical-related technological dependence, asthma, reactive airway or other chronic respiratory disease that requires daily medication for control.

Two MAB therapies were offered at Northwell, based upon availability: bamlanivimab (Eli Lilly and Company) and casirivimab/imdevimab (Regeneron Pharmaceuticals, Inc.).

### Monoclonal Antibody Infusion Operations

Northwell established a taskforce of clinicians paired with an operational team to develop a four-phase strategy and operational plan for MAB infusion. The initial phase established five outpatient infusion sites, all located on hospital campuses in free-standing buildings or in mobile hospital tents previously erected to accommodate the Spring 2020 COVID-19 surge.

In recognition that the emergency department (ED) is often the healthcare access point in underserved areas, phase 2 established MAB infusions directly for treat-and-release ED patients meeting EUA criteria, who otherwise lacked resources to travel to an infusion center. Phase 3 included administration of MAB therapy to eligible inpatients who developed COVID-19 while hospitalized for another cause and were COVID-19 negative on admission (all patients were tested upon hospital admission). The final phase included MAB therapy administration to patients in skilled nursing facilities, although with the rapid vaccine deployment supporting these facilities, this phase contributed only a small group of patients.

Information technology systems were configured to support patient referral, registration, and throughput in the infusion centers. Information about MAB therapy, the EUA, and referral instructions were disseminated widely to all Northwell’s New York metropolitan area affiliated providers. A dedicated call center and secure internal webpage were deployed to facilitate easy referral. The information collected included patient demographics and location; referring provider information; presence and details of COVID-19 symptoms and onset date; and screening of eligible comorbidities. The dedicated call center handled referrals, questions from providers and patients, and scheduling.

All patients were screened based on the EUA criteria at the time of referral. Infusion center staff training was created and deployed, including nursing competencies in biologic infusions as well as preparation with appropriate ACLS protocols and equipment in the event of an infusion reaction. Specific patient protocols were developed to treat patient reactions to the infusion, including rapid response team evaluation and transfer to a local ED most proximate to the infusion center, if necessary. To accommodate the EUA mandate for infusion within 10 days of COVID-19 symptom onset, the infusion centers were staffed 7 days a week.

### Study Population

All adult patients (age ≥18 years) who received MAB therapy in an ambulatory or ED location between November 20, 2020, and January 31, 2021, were included. Pediatric patients, inpatients, and skilled nursing facility patients that received MAB therapy in this date range were excluded from the analysis. Data collected include demographics, comorbidities, symptoms and their date of onset, date of COVID-19 test, and outcomes (including ED presentation, hospital admission, and mortality).

We further identified all patients age ≥18 years with a positive COVID-19 test between November 20, 2020, and January 31, 2021, who did not receive MAB therapy, but were eligible based upon EUA criteria. We excluded patients with a COVID-19 positive test or hospitalization prior to the study period. The outpatient outcomes of these patients are described, but not directly compared to the treatment group as we did not have symptoms (presence, timing, type, or severity) for the non-treatment group.

### Outcomes

We examined ED utilization and hospitalization within 28 days of a positive COVID-19 test for patients who received MAB therapy. Nine patients were missing a COVID-19 test date; for these patients we used the date of MAB therapy subtracted by the cohort median number of days from test to MAB therapy (4 days). The hospitalization rate by timing of MAB therapy relative to symptoms was also assessed.

For hospitalized patients, we performed a retrospective cohort study with a time-to-event survival analysis and a primary outcome of in hospital mortality. The control group was selected from the population of patients who met eligibility for MAB therapy, but did not receive it during the evaluation period.

### Covariates

We included sociodemographic and clinical features, including patient age, sex, race/ethnicity, number of hospital visits in the prior year, comorbidities, and presenting vital signs. Race/ethnicity was categorized as non-Hispanic White, non-Hispanic Black, Hispanic, Asian, Other/multiracial, and Unknown/declined. The comorbidities included DM, obesity, chronic respiratory conditions, COPD, CKD, hypertension, and immuno-compromising conditions (including the use of immunosuppressive medications). Presenting vital signs for patients hospitalized include heart rate, oxyhemoglobin saturation, temperature, and systolic and diastolic blood pressure.

### Statistical Analysis

We report descriptive statistics including median and interquartile range (IQR) for skewed continuous measures and proportions for categorical measures. We compared baseline clinical characteristics between patients who were and were not hospitalized using Fisher exact tests for categorical variables and nonparametric Kruskal-Wallis tests for continuous variables. Patients were categorized into 3 groups based upon timing of MAB therapy relative to symptom onset date (0-4 days, 5-7 days, and ≥8 days), to assess the difference in hospitalization rate.

For a univariable time-to-event analysis comparing mortality risk, we used the Kaplan-Meier survival curve to estimate in-hospital mortality to 28 days. Cox proportional-hazards regression models were used to estimate the association between MAB therapy and in-hospital mortality. We initially evaluated an unadjusted model, a model adjusted for age, sex, and race/ethnicity, and a model that added Charlson Comorbidity Index (CCI) to the prior model. The primary analysis used inverse probability weighting (IPW), whereby the predicted probabilities from a propensity-score model were used to calculate the stabilized IPW weight. The covariates included in the propensity model were age, sex, race/ethnicity, number of hospitalizations in prior year, and comorbidities and presenting vital signs (listed above, under Covariates).

All analyses were performed using the R programming language, version 4.0.3 (R Foundation for Statistical Computing, Vienna, Austria). A p-value <0.05 was considered significant. The Institutional Review Board of Northwell Health approved the study protocol before the commencement of the study. Individual-level informed consent was not obtained given the retrospective nature of the analysis of a large electronic medical record.

## RESULTS

From November 20, 2020, to January 31, 2021, 2818 adult patients with symptomatic COVID-19 received MAB infusion at Northwell Health: 2,745 (97%) ambulatory and 73 (3%) ED (Table 1). An additional 21 pediatric patients and 59 hospitalized patients received MAB therapy, and were not included in the analysis. The median patient age was 67 (IQR 58-74), and 59% were 65 years or older. Gender distribution was split evenly between males (1412 [50.1%]) and females (1406 [49.9%]). Most patients were non-Hispanic White (2061 [73%]), 104 (3.7%) were non-Hispanic Black and 168 (6%) were Hispanic. Hypertension was the most common comorbidity (1011 [36%]), followed by obesity (401 [23%]). The most common symptom was cough (seen in 1954 [70%] of patients), followed by malaise (1471 [53%]), fever (1422 [51%]), and headache (820 [30%]). While cough as the sole documented symptom was most common, many patients had multiple presenting symptoms (**eFigure 1 in the Supplement**).

**Table 1.** Characteristics of 2,818 patients with COVID-19 who received monoclonal antibody (MAB) therapy in ambulatory or emergency department setting.

|  | Overall<br>N=2818 | No Inpatient<br>Visit<br>N=2673 | Has Inpatient<br>Visit<br>N=145 | P-value |
| --- | --- | --- | --- | --- |
| Age (median [IQR]) | 67.00 [58.00,<br>74.00] | 66.00 [58.00,<br>74.00] | 75.00 [64.00,<br>82.00] | <0.001 |
| Age categories in years, N (%) |  |  |  | <0.001 |
| < 55 | 460 (16.3) | 450 (16.8) | 10 (6.9) |  |
| 55 – 64 | 710 (25.2) | 683 (25.6) | 27 (18.6) |  |
| 65 – 74 | 964 (34.2) | 930 (34.8) | 34 (23.4) |  |
| 75 + | 684 (24.3) | 610 (22.8) | 74 (51.0) |  |
| Female sex, N (%) | 1406 (49.9) | 1343 (50.2) | 63 (43.4) | 0.131 |
| Race/ethnicity, N (%) |  |  |  | 0.39 |
| Hispanic | 168 (6.0) | 160 (6.0) | 8 (5.5) |  |
| Non-Hispanic Black | 104 (3.7) | 99 (3.7) | 5 (3.4) |  |
| Asian | 110 (3.9) | 103 (3.9) | 7 (4.8) |  |
| Non-Hispanic White | 2061 (73.1) | 1948 (72.9) | 113 (77.9) |  |
| Other/multiracial | 332 (11.8) | 323 (12.1) | 9 (6.2) |  |
| Unknown/Declined | 43 (1.5) | 40 (1.5) | 3 (2.1) |  |
| Comorbidities, N (%) |  |  |  |  |
| Obesity | 401 (23.3) | 377 (23.4) | 24 (21.4) | 0.712 |
| Diabetes mellitus | 484 (17.2) | 421 (15.8) | 63 (43.4) | <0.001 |
| Hypertension | 1011 (35.9) | 901 (33.7) | 110 (75.9) | <0.001 |
| Chronic kidney disease | 113 (4.0) | 85 (3.2) | 28 (19.3) | <0.001 |
| Chronic obstructive pulmonary disease | 434 (15.4) | 394 (14.7) | 40 (27.6) | <0.001 |
| Chronic respiratory disease | 463 (16.4) | 418 (15.6) | 45 (31.0) | <0.001 |
| Immunosuppressed | 179 (6.4) | 161 (6.0) | 18 (12.4) | 0.004 |
| COVID-19 Symptoms |  |  |  |  |
| Cough | 1954 (70.4) | 1847 (70.2) | 107 (74.3) | 0.339 |
| Malaise | 1471 (53.0) | 1398 (53.1) | 73 (50.7) | 0.627 |
| Fever | 1422 (51.2) | 1350 (51.3) | 72 (50.0) | 0.825 |
| Headache | 820 (29.5) | 788 (30.0) | 32 (22.2) | 0.059 |
| Sore throat | 555 (20.0) | 532 (20.2) | 23 (16.0) | 0.257 |
| Gastrointestinal | 371 (13.4) | 351 (13.3) | 20 (13.9) | 0.95 |
| Loss taste/smell | 309 (11.1) | 296 (11.3) | 13 (9.0) | 0.49 |
| Muscle pain | 256 (9.2) | 240 (9.1) | 16 (11.1) | 0.512 |
| Shortness of breath | 143 (5.2) | 131 (5.0) | 12 (8.3) | 0.114 |
| Monoclonal Antibody Type, N |  |  |  | 0.304 |
| (%) |  |  |  |  |
| Casirivimab/<br>imdevimab | 317 (11.2) | 305 (11.4) | 12 (8.3) |  |
| Bamlanivimab | 2501 (88.8) | 2368 (88.6) | 133 (91.7) |  |
| Monoclonal Antibody Timing,<br>median (IQR) |  |  |  |  |
| Days from symptom<br>onset to therapy | 6.00 [4.00,<br>8.00] | 6.00 [4.00,<br>8.00] | 6.00 [5.00,<br>8.00] | 0.391 |
| Days from symptom<br>onset to COVID-19<br>test | 2.00 [1.00,<br>3.00] | 2.00 [1.00,<br>3.00] | 2.00 [1.00,<br>3.25] | 0.026 |
| Emergency department and<br>hospital utilization |  |  |  |  |
| ED Visit within 28<br>days, N (%) | 123 (4.4) | 112 (4.2) | 11 (7.6) | 0.082 |
| Days from COVID-19<br>test to ED visit<br>(median [IQR]) | 7.00 [5.00,<br>11.00] | 7.00 [5.00,<br>11.00] | 6.00 [3.00,<br>10.50] | 0.494 |
| Days from therapy to<br>ED visit (median [IQR]) | 3.00 [0.00,<br>6.00] | 3.00 [0.00,<br>6.00] | 2.00 [0.50,<br>4.50] | 0.558 |
| Days from COVID-19<br>test to hospitalization<br>(median [IQR]) |  |  | 7.00 [5.00,<br>11.00] | NA |
| Days From therapy to<br>hospitalization<br>(median [IQR]) |  |  | 3.00 [1.00,<br>8.00] | NA |

Most patients developed symptoms prior to COVID-19 test (median 2 days [IQR 1-3]; **eFigure 2 in the Supplement**). Among the patients with known symptom onset date, the median time from symptom onset to MAB therapy was 6 days (IQR 4-8; **eFigure 3 in the Supplement**). MAB referral to infusion scheduling occurred in under half a day (median 0.05 days [IQR 0.01-0.54]), and the MAB infusion occurred a median 1.75 days after referral (IQR 0.85-1.88). Most patients received bamlanivimab (2501 [89%]), with the remainder receiving casirivimab/imdevimab (317 [11%]).

Following MAB therapy and within 28 days of COVID-19 test, 123 patients (4.4%) presented to the ED and were released, a median of 7 days (IQR 5-11) from COVID-19 test. In a similar timeframe, 145 patients (5.1%) who received MAB therapy were hospitalized, a median of 7 days (IQR 5-11) after COVID-19 test. The median time from MAB therapy to ED presentation therapy was 3 days (IQR 0-6), and the median time from MAB therapy to hospitalization was 3 days (IQR 1-8). A greater proportion of patients who were hospitalized following MAB therapy had comorbidities, including diabetes, hypertension, chronic kidney disease, respiratory disease, and immunosuppressive disease (see **Table 1**).

In the subgroup of patients where symptom onset date was known (2,721 [96.6%]), the hospitalization rate within 28 days of COVID-19 test was 4.4% (95% confidence interval [95% CI] 2.9-5.9%) for patients who received MAB therapy early (within 0-4 days of symptom onset), 5% (95% CI 3.6-6.2%) for those within 5-7 days, and 6.1% (95% CI 4.6-7.4%) for those who received it ≥8 days, although this was not statistically significant (p-value for trend = 0.15; **Figure 1**).

**Figure 1.**
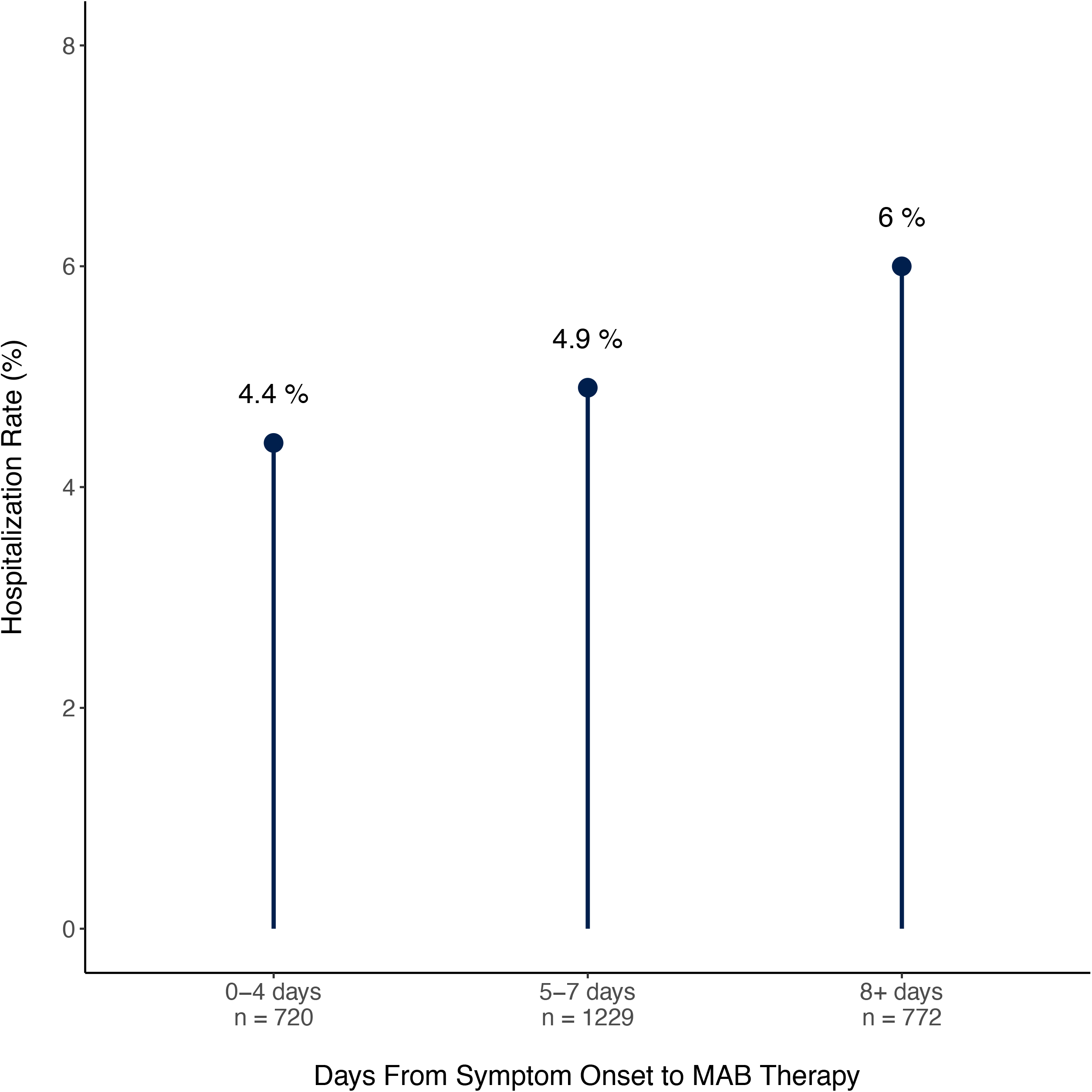
Timing of monoclonal antibody (MAB) therapy and hospitalization rate. Among 2,721 patients with known symptom onset date, the hospitalization rate within 28 days of COVID-19 test was 4.4% for patients who received MAB therapy early (within 0-4 days of symptom onset), 5% for those in a middle group (5-7 days), and 6.1% for those who received it late (≥8 days).

Among 2,713 COVID-19 positive patients meeting eligibility criteria based upon age or comorbidities but not receiving MAB therapy, the median age was 66 years old (IQR 55-73) and 55% were female. Non-Hispanic White patients were most common (1,596 [59%]), and there were 183 (6.7%) non-Hispanic Black and 334 (12.3%) Hispanic patients. Symptoms were not ascertained for this group, but similar to the MAB therapy group, hypertension was the most common comorbidity (1119 [41%]). A total of 142 patients (5.2%) and 200 (7.4%) of patients in this group had an ED visit and inpatient hospitalization, respectively, within 28 days of COVID-19 test. Patients hospitalized had a higher burden of comorbid conditions (**eTable in the Supplement**).

### Hospital Outcomes

A total of 145 MAB patients were hospitalized and were compared with 200 controls who otherwise met MAB therapy eligibility criteria and were hospitalized (**Table 2**). The MAB group was slightly older (median age 75 years [IQR 64-82] vs. 69 [57-78]), with a lower proportion of women (43% vs. 53%), and a higher proportion of non-Hispanic White race (78% vs. 59%). There was no significant difference in the presence of comorbidities between the groups.

**Table 2.** Characteristics of patients who received and did not received pre-hospital monoclonal antibody (MAB) therapy and were hospitalized within 28 days of COVID-19 test.

|  | All<br>Hospitalized<br>Patients<br>N=345 | Control Group<br>N=200 | Monoclonal<br>Antibody<br>Treatment<br>Group<br>N=145 | P-value |
| --- | --- | --- | --- | --- |
| n | 345 | 200 | 145 |  |
| Age (median [IQR]) | 72.00 [61.00,<br>80.00] | 69.00 [57.00,<br>78.00] | 75.00 [64.00,<br>82.00] | 0.001 |
| Age categories in years, N (%) |  |  |  | 0.001 |
| < 55 | 52 (15.1) | 42 (21.0) | 10 (6.9) |  |
| 55 – 64 | 62 (18.0) | 35 (17.5) | 27 (18.6) |  |
| 65 – 74 | 89 (25.8) | 55 (27.5) | 34 (23.4) |  |
| 75 + | 142 (41.2) | 68 (34.0) | 74 (51.0) |  |
| Female sex, N (%) | 169 (49.0) | 106 (53.0) | 63 (43.4) | 0.1 |
| Race/ethnicity, N (%) |  |  |  | 0.02 |
| Hispanic | 32 (9.3) | 24 (12.0) | 8 (5.5) |  |
| Non-Hispanic Black | 25 (7.2) | 20 (10.0) | 5 (3.4) |  |
| Asian | 19 (5.5) | 12 (6.0) | 7 (4.8) |  |
| Non-Hispanic White | 231 (67.0) | 118 (59.0) | 113 (77.9) |  |
| Other/multiracial | 29 (8.4) | 20 (10.0) | 9 (6.2) |  |
| Unknown/Declined | 9 (2.6) | 6 (3.0) | 3 (2.1) |  |
| Comorbidities, N (%) |  |  |  |  |
| Obesity | 73 (23.4) | 49 (24.5) | 24 (21.4) | 0.635 |
| Diabetes mellitus | 149 (43.2) | 86 (43.0) | 63 (43.4) | 1 |
| Hypertension | 259 (75.1) | 149 (74.5) | 110 (75.9) | 0.871 |
| Chronic kidney disease | 50 (14.5) | 25 (12.5) | 25 (17.2) | 0.28 |
| Chronic obstructive<br>pulmonary disease | 95 (27.5) | 55 (27.5) | 40 (27.6) | 1 |
| Chronic respiratory disease | 113 (32.8) | 68 (34.0) | 45 (31.0) | 0.643 |
| Immunosuppressed | 38 (11.0) | 20 (10.0) | 18 (12.4) | 0.594 |
| Charlson Comorbidity Index (median<br>[IQR]) | 6.00 [4.00,<br>8.00] | 5.00 [3.00,<br>8.00] | 6.00 [4.00,<br>8.00] | 0.219 |
| Presentation vital signs, median<br>(IQR) |  |  |  |  |
| Heart rate, beats per minute | 89.00 [78.00,<br>102.00] | 89.50 [77.00,<br>103.00] | 89.00 [79.00,<br>100.00] | 0.575 |
| Systolic blood pressure,<br>mmHg | 131.00<br>[119.00,<br>147.00] | 131.50<br>[118.75,<br>146.25] | 130.00<br>[121.00,<br>147.00] | 0.981 |
| Diastolic blood pressure, mmHg | 74.00 [67.00, 83.00] | 74.00 [67.00, 83.00] | 75.00 [66.00, 82.00] | 0.435 |
| Temperature, degrees Celsius | 37.00 [36.70, 37.60] | 36.90 [36.70, 37.42] | 37.10 [36.70, 37.70] | 0.036 |
| Oxygen saturation, % | 96.00 [92.00, 98.00] | 96.00 [92.00, 98.00] | 96.00 [93.00, 97.00] | 0.478 |

In the MAB group, 16 (11%) patients met the primary outcome of in-hospital mortality, versus 21 (10.5%) in the control group. Kaplan Meier survival curve showed no difference between the two groups for event-free probability (log rank p-value = 0.41; **Figure 2**). In an unadjusted Cox proportional hazards model, the hazard ratio (HR) for time to inpatient mortality for the MAB group was 1.38 (95% CI 0.696-2.719). There was no significant association between pre-hospitalization MAB use and the primary end point in both a model adjusted for demographics (HR 1.1, 95% CI 0.53-2.23), a model adjusted for demographics and CCI (HR 1.22, 95% CI 0.573-2.59), and a model with inverse probability weighting according to the propensity score (HR 1.19, 95% CI 0.619-2.29).

**Figure 2.**
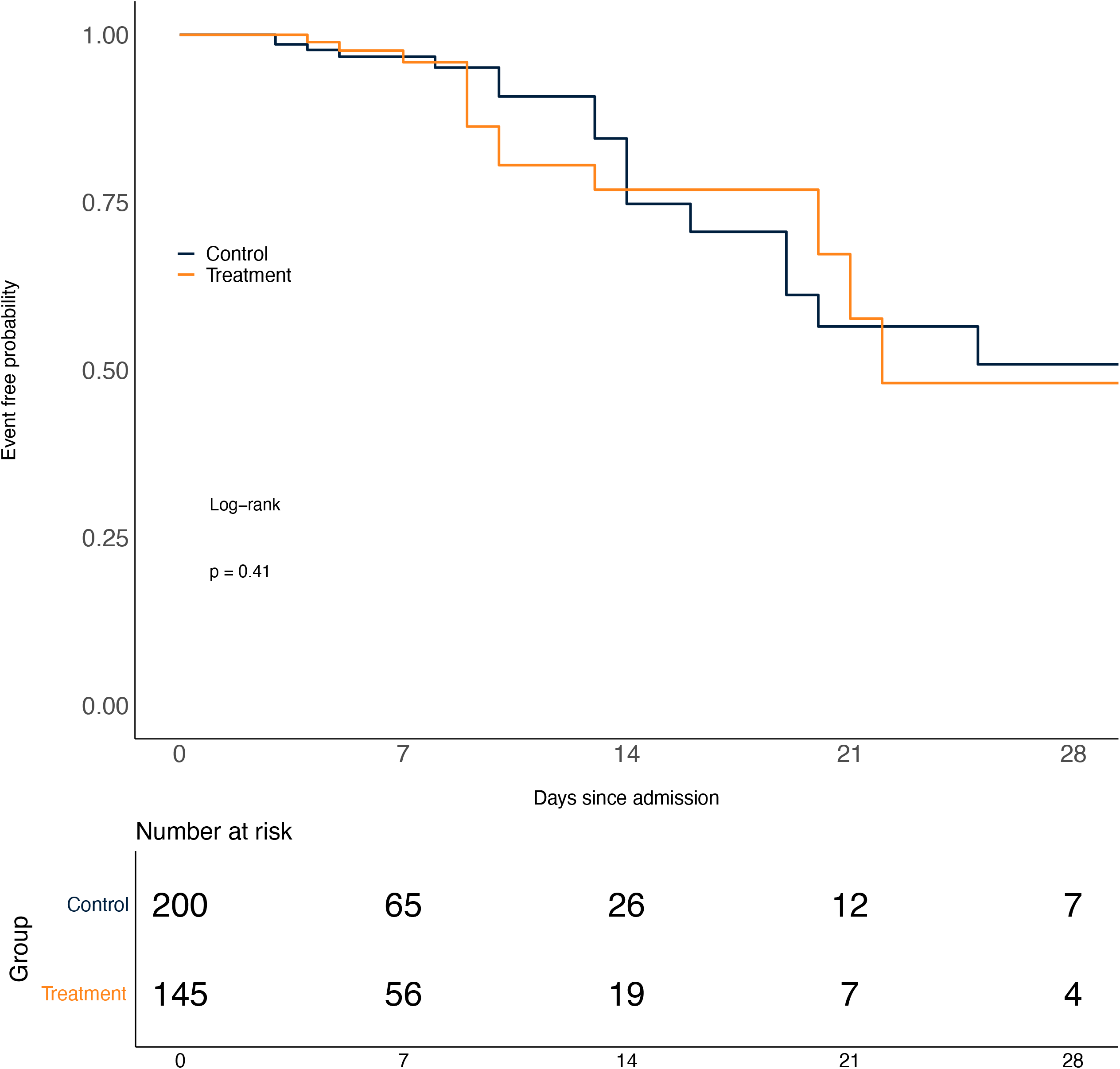
Freedom from end point of in-hospital mortality.

## DISCUSSION

Despite the issuance of an FDA EUA for two MAB therapies in late 2020 to treat mild to moderate COVID-19 in high-risk outpatients, adoption and utilization nationally has been slow.^5^ Hesitancy may be related to questions of treatment effectiveness, logistical challenges, and staffing requirements during the pandemic.^9^ In the 2.5 month period following the EUA, Northwell scaled up an ambulatory MAB infusion operation and successfully administered therapy to over 2,800 eligible patients with most patients receiving therapy within 1.8 days of referral. The operational success required close collaboration and coordination of clinical, operation, informatics, information technology, ambulatory, and population health leadership to ensure the appropriate requirements were in place.

Among the patients who received MAB therapy, a majority received bamlanivimab due to availability. A total of 145 patients (5.1%) were hospitalized within 28 days of COVID-19 test, and 16 died (0.6% of total population and 11% of hospitalized patients). We did find a trend toward a lower rate of hospitalization for patients receiving therapy more proximate to symptom onset date, although this finding was not statistically significant. Inasmuch as the effect of MAB therapy is to reduce SARS-CoV-2 viral load,^11^ receiving these therapies earlier in the disease course should be beneficial; the low numbers of hospitalized patients in our treatment group may contribute to the lack of statistical significance. Among the 2,713 patients who tested positive for COVID-19 during the same time period in our health system, and who met age or comorbidity eligibility criteria for MAB yet did not receive it, 200 (7.4%) were hospitalized within 28 days. A direct comparison to our MAB cohort, however, is not feasible given the lack of symptom data for these non-MAB patients.

Compared to a matched control group, there was no significant difference in the hospital outcome of in-hospital mortality. While our sample size of hospitalized patients is small, this finding may be more related to COVID-19 disease burden: once a patient meets clinical requirements for hospitalization, prior MAB therapy likely does not alter the clinical trajectory. Indeed, randomized trials of MAB in hospitalized patients did not demonstrate efficacy ^4^.

While the published randomized control trials to date presented promising efficacy data, the primary endpoint was focused on viral load rather than clinically meaningful outcomes such as hospitalization or death.^2,3^ A case series suggesting benefit has been described, but suffers from small sample size and lack of control.^12^ We were able to describe the outcomes in 2,818 patients receiving MAB therapy and further compared in-hospital mortality with an appropriately matched control group. Our study did not demonstrate effectiveness of MAB therapy on preventing in-hospital mortality and we did not have a control group to examine the effectiveness of MAB therapy on preventing hospitalization. Nonetheless, the trend toward reduced hospitalization seen in the early treatment cohort is intriguing: timely referral and operational efficiency to administer MAB therapy early in the course of disease would benefit hospital operations by reducing the burden on capacity issues. Although we invested resources to specifically staff the MAB infusion facilities, such a derived benefit may outweigh the MAB resource utilization. Certainly, preventing mortality is the most critical outcome, however, a reduced burden of critically ill patients would allow the hospital staff to focus on non-COVID-19 patients as well. In addition, the administration of MAB therapy in the ED helped facilitate health equity, since many underserved communities, challenged by the lack of primary care and a high prevalence of comorbid conditions, were disproportionately affected by severe COVID-19 disease. Interestingly, this phase of our MAB program did not result in ED overcrowding.

Future efforts for MAB therapy may include home infusion or mobile treatment options. Although these were considered in our original MAB strategy, staffing burden for the number of patients that could be treated was high and operational considerations such as preparation and transportation of the mixed MAB infusion were considerable. It is hoped that alternate routes (e.g., intramuscular or subcutaneous) of administering MAB therapies can be developed to offset these operational and staffing challenges.

As many health systems continue to deal with COVID-19 surges, we recommend establishing a national database to analyze MAB treatment in larger cohorts. While randomized, placebo-controlled trials may not be logistically feasible, further meta-analyses of centers leveraging these therapies may be in order.

### Limitations

Limitations of our study include the observational and retrospective study design. In addition, our health system is based in New York and may not be generalizable to other regions. Due to the lack of symptom documentation for our control group of patients, we were unable to assess the impact of MAB therapy on hospitalization rate. Given the small number of patients and low event rate, our analysis of inpatient mortality may be underpowered to detect a difference.

## Conclusions

The EUA for the MAB infusions provides a foundation for treatment of early mild to moderate COVID-19 in high-risk patients. This study describes the rapid development of a MAB infusion program to provide such treatment for over 2800 patients. Establishing the capability to perform MAB infusion therapy requires close collaboration and coordination of numerous stakeholders and can support hospital operations in the setting of a pandemic surge. Further investigation is required to define the optimal timing of MAB therapy and the potential attendant reduction in hospitalization and mortality.

## Data Availability

All data is available

## FIGURES

**eFigure 1. Documented symptoms for patients receiving monoclonal antibody (MAB) therapy**. The most common individual symptoms were cough, malaise, fever, and headache (horizontal bars, “Set size”). Many patients had multiple presenting symptoms (vertical bars, “Intersection size”).

**eFigure 2. Days from symptom onset to COVID-19 test**. The majority of patients had a COVID-19 test between 0 and 3 days (median 2 days [IQR 1-3]), although some patients had a test prior to symptom onset.

**eFigure 3. Days from symptom onset to monoclonal antibody (MAB) therapy**. The majority of patients received MAB therapy at 5-6 days after symptom onset (median 6 days [IQR 4-8]).

