## Supplemental Table 1 for "Early Experience With Neutralizing Monoclonal Antibody Therapy For COVID-19"

**Online Only Supplement Material**

**eTable 1. Characteristics of 2,713 patients with COVID-19 who did not receive monoclonal antibody (MAB) therapy.**

|  | Overall  N=2713 | No Inpatient Visit  N=2513 | Has Inpatient Visit  N=200 | P-value |
| --- | --- | --- | --- | --- |
| Age (median [IQR]) | 66.00 [55.00, 73.00] | 66.00 [55.00, 73.00] | 69.00 [57.00, 78.00] | <0.001 |
| AgeGroup (%) |  |  |  | <0.001 |
| < 55 | 644 (23.7) | 602 (24.0) | 42 ( 21.0) |  |
| 55 - 64 | 576 (21.2) | 541 (21.5) | 35 ( 17.5) |  |
| 65 - 74 | 926 (34.1) | 871 (34.7) | 55 ( 27.5) |  |
| 75 + | 567 (20.9) | 499 (19.9) | 68 ( 34.0) |  |
| Female sex, N (%) | 1497 (55.2) | 1391 (55.4) | 106 ( 53.0) | 0.569 |
| Race/ethnicity, N (%) |  |  |  | <0.001 |
| Hispanic | 335 (12.3) | 311 (12.4) | 24 (12.0) |  |
| Non-Hispanic Black | 183 ( 6.7) | 163 ( 6.5) | 20 (10.0) |  |
| Asian | 69 ( 2.5) | 57 ( 2.3) | 12 ( 6.0) |  |
| Non-Hispanic White | 1596 (58.8) | 1478 (58.8) | 118 (59.0) |  |
| Other/multiracial | 191 ( 7.0) | 171 ( 6.8) | 20 (10.0) |  |
| Unknown/Declined | 339 (12.5) | 333 (13.3) | 6 ( 3.0) |  |
| Comorbidities, N (%) |  |  |  |  |
| Diabetes mellitus | 523 (19.3) | 437 (17.4) | 86 ( 43.0) | <0.001 |
| Hypertension | 1119 (41.2) | 970 (38.6) | 149 ( 74.5) | <0.001 |
| Chronic kidney disease | 112 ( 4.1) | 85 ( 3.4) | 27 ( 13.5) | <0.001 |
| Chronic obstructive pulmonary disease | 435 (16.0) | 380 (15.1) | 55 ( 27.5) | <0.001 |
| Chronic respiratory disease | 471 (17.4) | 403 (16.0) | 68 ( 34.0) | <0.001 |
| Immunosuppressed | 315 (11.6) | 295 (11.7) | 20 ( 10.0) | 0.533 |
| Emergency department and hospital utilization |  |  |  |  |
| ED Visit within 28 days, N (%) | 142 ( 5.2) | 112 ( 4.5) | 30 ( 15.0) | <0.001 |
| Days from COVID-19 test to ED visit (median [IQR]) | 8.00 [4.00, 12.00] | 8.00 [4.00, 12.00] | 6.00 [3.00, 13.75] | 0.306 |
| Days from COVID-19 test to hospitalization (median [IQR]) |  |  | 7.00 [3.00, 11.00] |  |

**eFigure 1. Documented symptoms for patients receiving monoclonal antibody (MAB) therapy.** The most common individual symptoms were cough, malaise, fever, and headache (horizontal bars, "Set size"). Many patients had multiple presenting symptoms (vertical bars, "Intersection size").

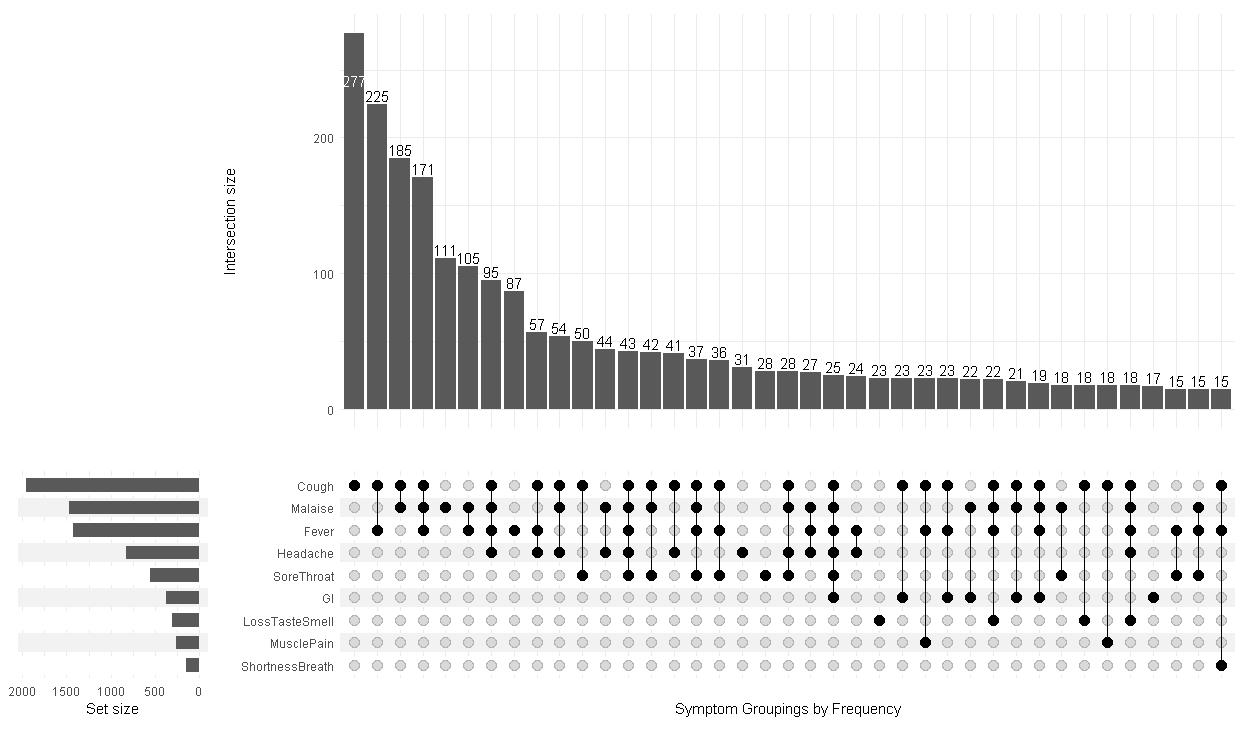

**eFigure 2. Days from symptom onset to COVID-19 test.** The majority of patients had a COVID-19 test between 0 and 3 days (median 2 days [IQR 1-3]), although some patients had a test prior to symptom onset.

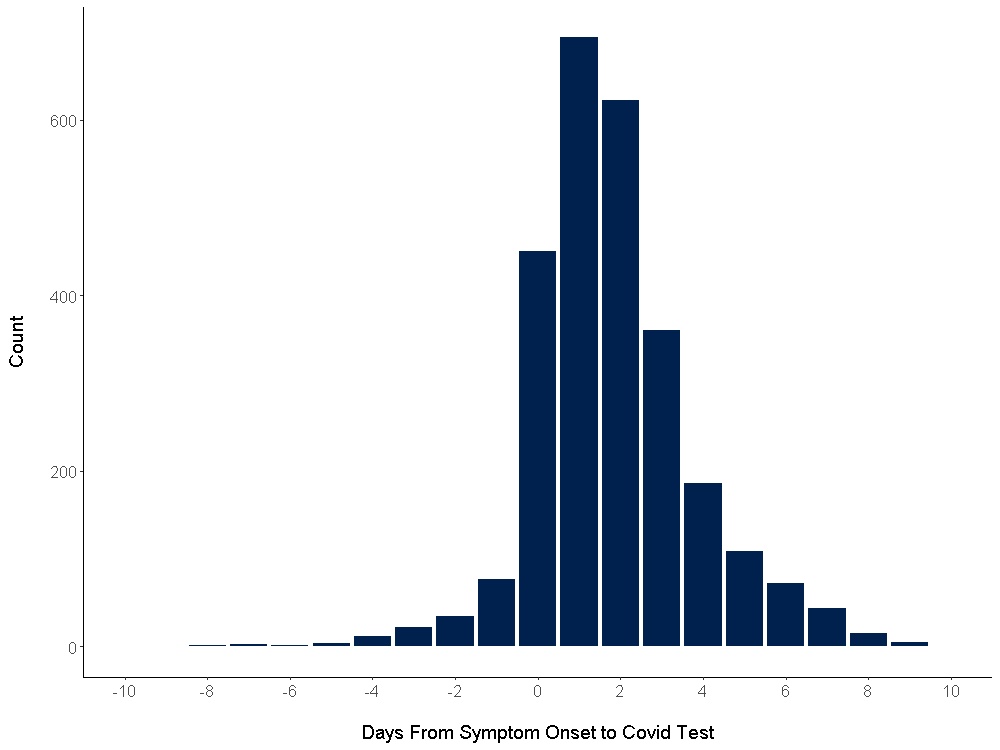

**eFigure 3. Days from symptom onset to monoclonal antibody (MAB) therapy.** The majority of patients received MAB therapy at 5-6 days after symptom onset (median 6 days [IQR 4-8]).

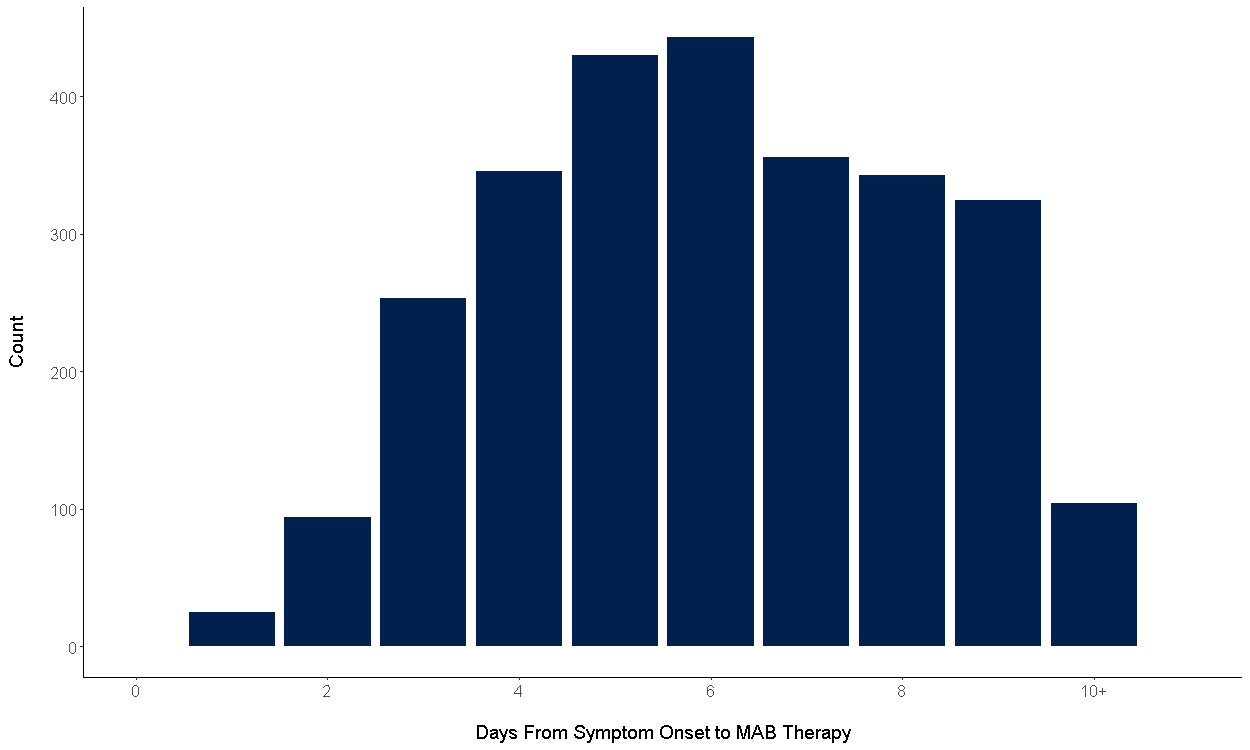
